# Sex-specific Trends in the Use of Temporary Mechanical Circulatory Support in patients listed for Orthotopic Heart Transplant Before and After the UNOS Allocation System Change

**DOI:** 10.1101/2025.02.11.25322110

**Authors:** Nicole Cyrille-Superville, Priyesh A Patel, Brian N White, Snehal R Patel, Rachel Garcia, Heather Rose, Susan Bernado, Lauren Harmon, Shuktika Nandkeolyar, Joseph D Mishkin, Sanjeev K Gulati, Amar Doshi, Theodore A Frank, Adam D Devore

**Affiliations:** Sanger Heart and Vascular Institute, Atrium Health, Charlotte NC; Wake Forest University School of Medicine; Department of Medicine, Division of Cardiology, Montefiore Medical Center, Albert Einstein College of Medicine, Bronx, NY; Department of Medicine, Duke University School of Medicine, Durham, NC

## Abstract

**Background:** Since the United Network for Organ Sharing (UNOS) allocation system change patients listed for orthotopic heart transplant (OHT) are more likely to be on temporary mechanical circulatory support (tMCS). Limited sex specific data exits for use and outcomes of tMCS since the allocation change.

**Methods:** We queried the UNOS registry for patients listed for OHT on tMCS from October 1, 2015 to June 28, 2023, comparing baseline characteristics and outcomes between sexes pre- and post-allocation change.

**Results:** Women comprised 23% of patients listed for OHT on tMCS before and after the allocation change, despite similar cardiac index (CI) compared with men (pre: 2.03 vs 2.09 L/min m^2^ p=0.21; post: 1.92 vs 1.92 L/min m^2^ p=0.89). Women were significantly younger (54 vs 57 years; p<0.001), had lower BMI (26.9 vs 27.2; p 0.006), were more likely to be on intra-aortic balloon pump (IABP) (67% vs 61% p <0.001) and had shorter waitlist times (13 vs 16 days; p <0.001). Waitlist mortality decreased similarly for both sexes (3.6% vs 3.8%; p= 0.7). There was no significant difference in 1-year post-transplant survival between the sexes in either era (HR: 0.9, 95% CI: 0.76, 1.06 p=0.2), however, 1-year post-transplant survival improved overall for patients bridged with tMCS post allocation change independent of sex (HR 0.70 (95% CI: 0.54 to 0.90; p=0.005).

**Conclusion:** Despite an increase in the use of tMCS as a bridge to OHT since the UNOS allocation change, women comprise less than a quarter of patients listed on tMCS despite similar CI, suggesting possible underuse in women. Notably, for those bridged to OHT in the post allocation era, waitlist mortality has improved for both sexes and 1-year post-transplant survival has also improved independent of sex.

## Introduction

In 2018, the United Network of Organ Sharing (UNOS) allocation system was changed to prioritize patients listed with the greatest clinical urgency for orthotopic heart transplant (OHT), whereby the previous single highest urgency status (1A) was broken down into 3 separately ranked statuses in descending order of illness (new status 1, 2, and 3). Patients in cardiogenic shock supported with extracorporeal membrane oxygenation (ECMO) or other non-dischargeable biventricular mechanical circulatory support (MCS) are assigned to the highest urgency status and those with lesser degrees of support are distributed into a descending rank order of priority. **(1,2)** Since the allocation change, there has been a significant increase in patients on temporary MCS (tMCS) listed for OHT, with a simultaneous decrease in patients bridged with durable left ventricular assist devices (LVAD). **(3)**

Prior studies have highlighted sex disparities among patients receiving durable LVAD, with women on LVAD support less likely to receive OHT and more likely to experience waitlist removal and waitlist death. **(4,5)** Moreover, in cardiogenic shock women have been shown less likely to receive acute tMCS compared with their male counterparts. **(6,7)** We therefore sought to investigate the UNOS registry for sex specific trends in the use and outcomes of tMCS among patients listed for OHT, before and after the UNOS allocation change.

### Methods

A retrospective analysis of the combined UNOS thoracic database was performed. This publicly available data contains no patient identifiers and as such was deemed exempt from Institutional Review Board review. The database contains information on all waitlist registrations and OHT recipients listed or performed in the United States and reported to the Organ Procurement and Transplantation Record since October 1, 1987. The database was queried for all adults (aged ≥18 years) listed for OHT between October 1, 2015 and June 28, 2023. Only patients on tMCS at the time of OHT listing were included in this analysis. Patients with a history of previous transplant and those listed for multiorgan transplants were excluded. Era 1 was defined as pre-allocation change (October 1, 2015−October 17, 2018), and era 2 was defined as post-allocation change (October 18, 2018−June 28, 2023). The baseline characteristics and outcomes of patients listed on tMCS in eras 1 and 2 were compared. The model for transplantation rate was fit on the full cohort, which included any patient on the OHT waitlist who satisfied the inclusion/exclusion criteria discussed above. In contrast, the models for post-transplant survival were fit on subsets of the data that included only those patients who received OHT, while waitlist-survival included only those who did not receive OHT.

### Statistical Analysis

The statistical analysis was performed on processed thoracic national STAR (Standard Transplant and Research) files obtained from UNOS. Descriptive statistics and tests of association including Fisher’s exact test, Pearson’s chi-squared test, and Wilcoxon-rank sum test where appropriate were calculated for relevant patient characteristics, stratified by UNOS allocation period and sex. Kaplan-Meier survival curves for post-transplant and waitlist outcomes were generated and stratified by UNOS allocation period and sex. The log-rank test was applied to compare unadjusted differences between these stratified groups. Multivariate Cox proportional hazards models were employed to evaluate the impact of the UNOS allocation period on the effect of sex on both post-transplant survival and waitlist survival, adjusting for relevant patient characteristics such as medical history and demographic information. The proportional hazards assumption was checked for all models and not found to be violated. Similarly, a multivariate logistic regression model was used to assess whether the UNOS allocation period influenced the effect of sex on the overall transplantation rate, with adjustments for relevant patient characteristics. All analyses were performed in R version 4.1.1 (2021-08-10).

## Results

### Sex-specific Trends in the use of tMCS

From Oct 2015 to June 2023 there was a notable surge in the overall use of tMCS as a bridge to OHT among both men and women (Figure 1). Among women, tMCS use increased from approximately 8.4% to 36% and for men from 11% to 42%. Women comprised 23% of patients listed for OHT on tMCS, both before and after the allocation change. A significantly higher proportion of women on tMCS underwent OHT compared with men before the allocation change (74% vs 66%, p=0.026). However, post allocation change, the frequency of OHT was similar and higher for both women and men on tMCS, 87% and 89% respectively (Table 1). Women in the post allocation change period were more likely to be bridged via intra-aortic balloon pump (IABP) compared with men (67% vs 61% p <0.001) and have lower use of non-dischargeable MCS (19% vs 28%, p<0.001). The use of ECMO has increased for both men (pre 1.1% vs post 10%) and women (pre 0.5% vs post 13%). On the contrary, the utilization of TAH has markedly decreased since the allocation change for both sexes (men: pre 14% vs post 0.9%; women: pre 5.9% vs post 0.6%). The majority of men and women listed on tMCS post allocation change were listed status 2, though women less so than men (81% vs 84%; p 0.004).

**TABLE 1.**
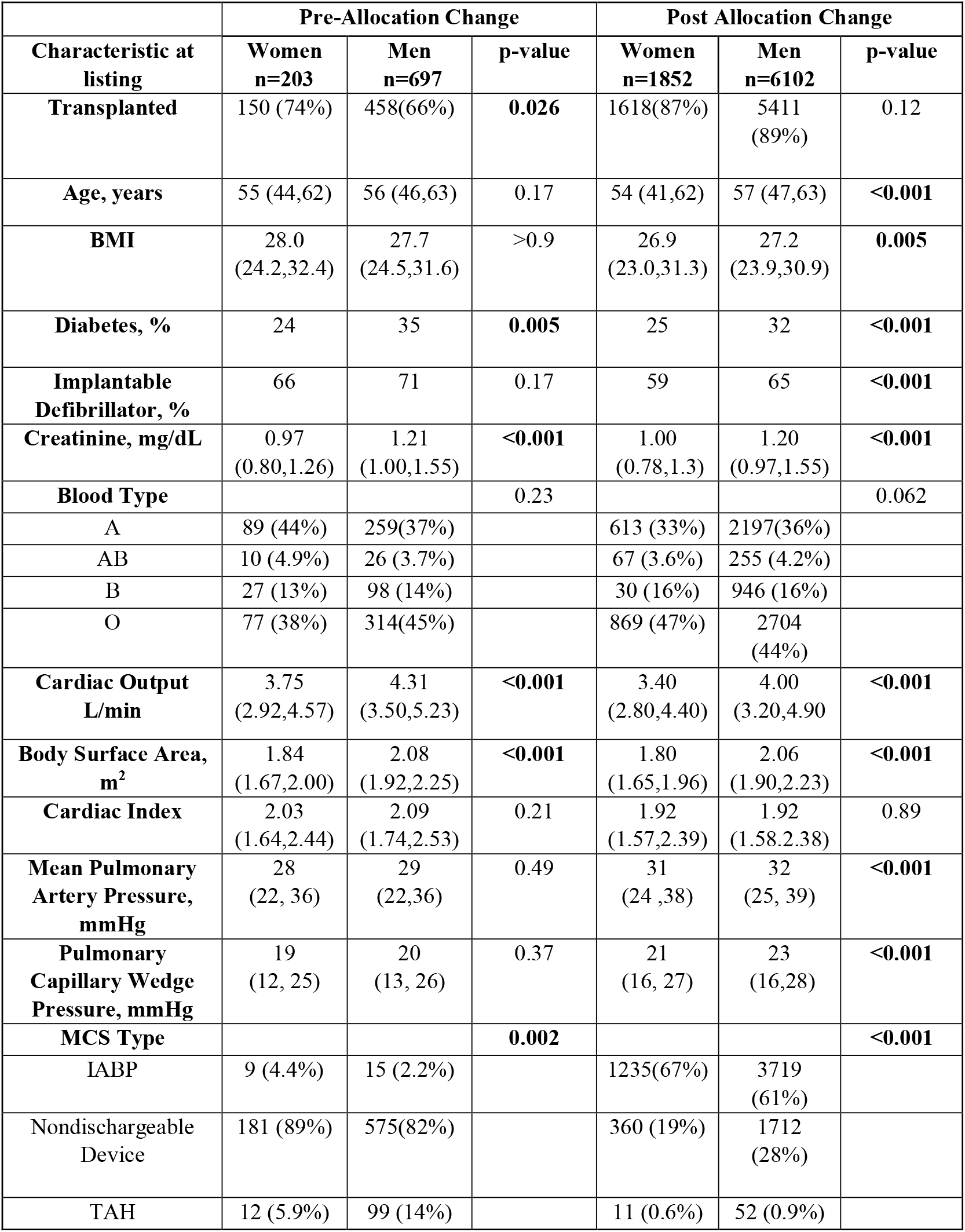
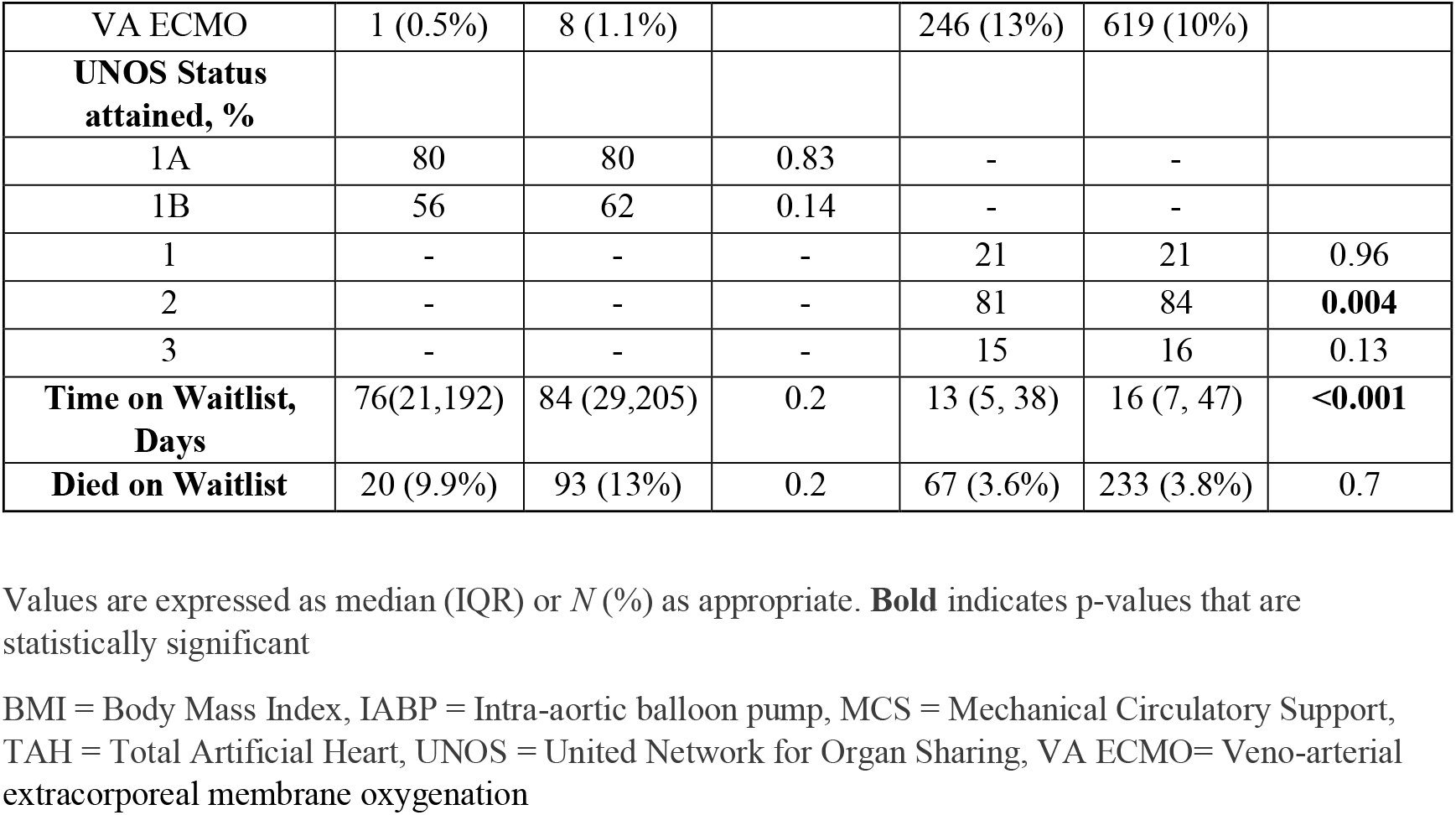
Baseline Characteristics and Outcomes for Patients Listed for Orthotopic Heart Transplant on Temporary Mechanical Circulatory Support.

**Figure 1.**
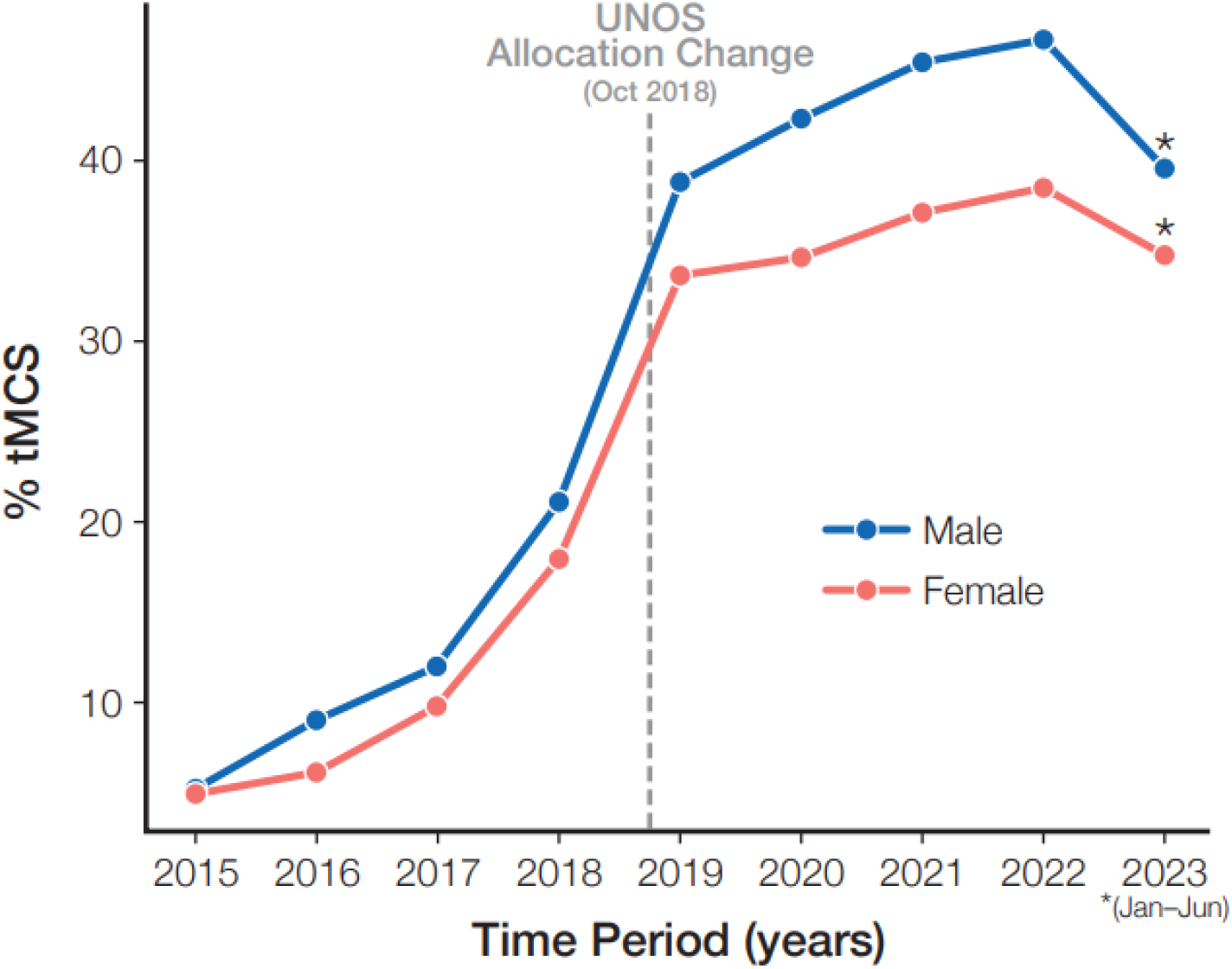
National Trend in the Use of Temporary Mechanical Circulatory Among Men and women.

**Figure 2.**
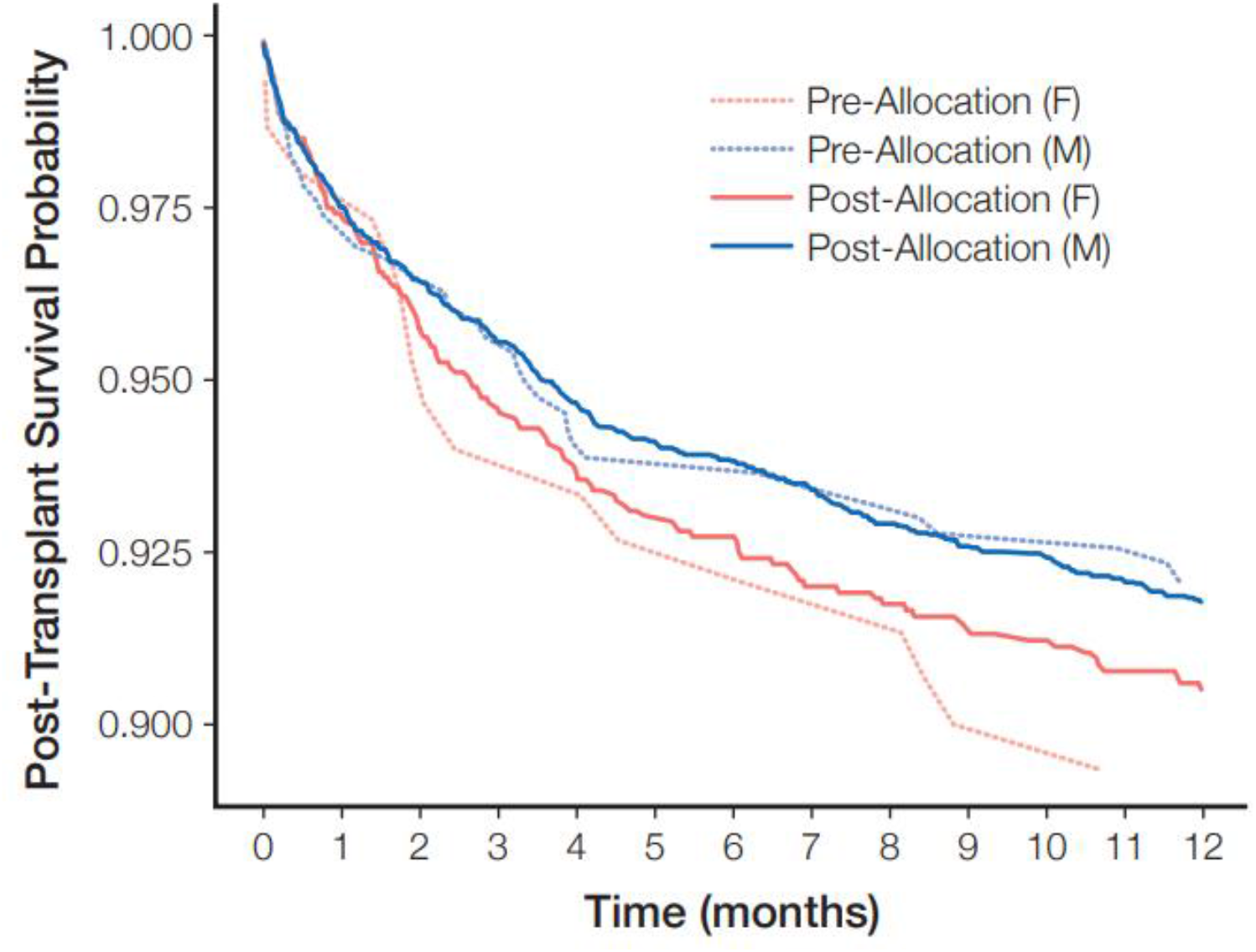
One-year Post Transplant Survival Before and After UNOS Allocation Change stratified by Sex.

### Sex-Related Clinical Differences

Baseline characteristics of patients at time of listing are illustrated in Table 1. Post allocation change, women on tMCS were significantly younger (54 vs 57 years; p<0.001), with lower BMI (26.9 vs 27.2; p 0.006) and less likely to have an implantable defibrillator (59% vs 65%; p<0.001) compared with men. In both the pre- and post-allocation periods, women were less likely to be diabetic. Before the allocation change, filling pressures including mean pulmonary artery pressures (PAPm) and pulmonary capillary wedge pressure (PCWP) were similar between the sexes. However, in the post allocation change era filling pressures including PAPm (31mmHg vs 32mmHg p< 0.001) and PCWP (21 mmHg vs 23 mmHg p <0.001), were significantly lower in women at the time of listing. Women had significantly lower cardiac output (CO) at time of listing pre- (3.75 vs 4.31 L/min p < 0.001) and post- (3.4 vs 4 L/min p <0.001) allocation change, though with similar cardiac index (pre: 2.03 vs 2.09 L/min m^2^ p=0.21, post: 1.92 vs 1.92 L/min m^2^ p=0.89).

### Trends in Outcomes

Time on the waitlist has decreased for both men (pre 84 vs post 16 days) and women (pre 76 vs post 13 days) from pre-to post-allocation change when bridged with tMCS. Women, however, have significantly shorter waitlist times compared with men post-allocation change (13 days vs 16 days; p <0.001) (Table 1). Waitlist mortality has improved post allocation change with no significant difference between women and men (pre: 9.9% vs 13% p = 0.2; post: 3.6% vs 3.8% p = 0.7). The 1-year post-transplant survival was similar between the sexes pre-and post-allocation change (HR: 0.9, 95% CI: 0.76, 1.06 p=0.2), however, overall 1-year post-transplant survival has improved for patients bridged with tMCS post allocation change independent of sex (HR 0.70 (95% CI: 0.54 to 0.90; p=0.005). Of note, when stratified by device type patients bridged with ECMO in the new era had significantly increased risk of death (HR: 1.44, 95% CI: 1.13 to 1.85 p=0.003) (Figure 3).

**Figure 3.**
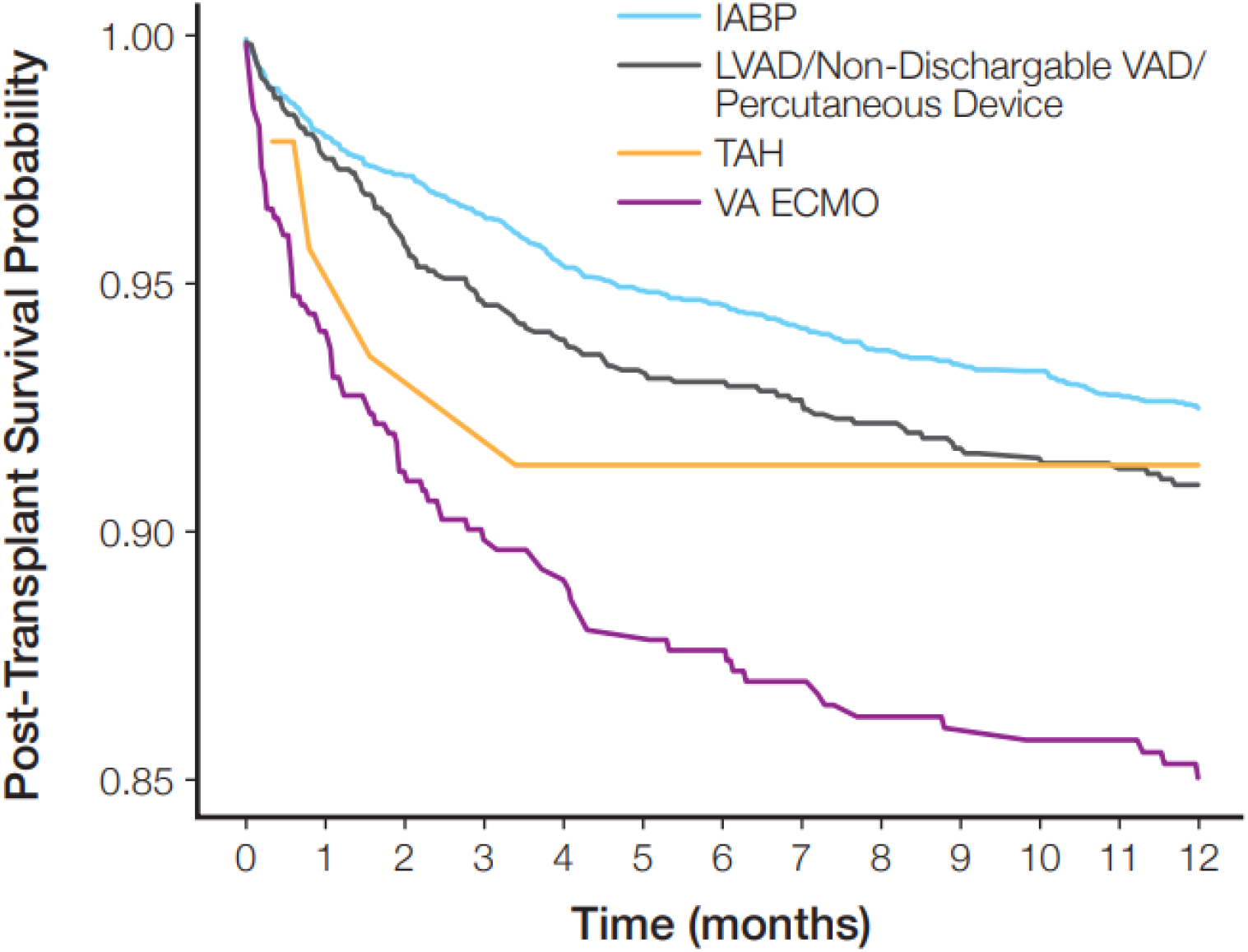
One-Year Post Transplant Survival in the Post-Allocation change Era Stratified by Device Type.

## Discussion

To our knowledge, this is the largest study to date examining sex specific trends and sex-related differences in the use and outcomes for patients placed on tMCS as bridge to OHT before and after the UNOS allocation change. Our major findings are as follows: 1) despite a marked increase in the use of tMCS since the allocation change, women still account for less than a quarter of those listed on tMCS despite similar CI at OHT listing; 2) women listed for OHT on tMCS before and after the allocation change are more likely to be bridged using IABP compared with men; 3) waitlist mortality has improved for both sexes; and 4) overall 1-year post-transplant survival has improved post allocation change for patients bridged with tMCS successfully to OHT independent of sex. Taken together our findings suggest that tMCS, though more frequently used since the allocation change with similar outcomes between the sexes, may still be underused in women as a bridge to OHT compared with their male counterparts.

Potential bias for underutilization for heart failure therapies in women has been previously reported. For instance, as demonstrated in prior studies, women in our cohort were less likely to have a defibrillator at the time of listing compared with men. **(8-10)** Our findings are also consistent with prior studies showing the use of tMCS as well as durable LVAD are less often used in women, at times with resultant worse outcomes. **(4-7)** Vallabhajosyula et al, for example, found that in a cohort of more than 370,000 patients admitted with cardiogenic shock in the setting of acute myocardial infarction (AMI), men were significantly more likely to receive tMCS compared with women (50.4% vs 39.5% p <0.001), with higher inpatient mortality, palliative care consultation and Do Not Resuscitate status observed in women.**(6)**

UNOS data do not provide adequate fidelity to understand what patient features could have affected tMCS bridge strategies. However, women are generally smaller in size, supported by significantly lower BSA compared with men in this study (pre-AC 1.84 vs 2.08 p<0.001; post-AC 1.8 vs 2.06 p<0.001). It is possible smaller LV cavities and vasculature could have limited tMCS options due to concerns for vascular complications **(11,12)**. Furthermore, women are also more likely to be sensitized with pre-formed anti–human leukocyte antigen antibodies related to prior pregnancies, which could have limited tMCS options due to concern for bleeding and transfusion needs **(13,14)**. Indeed, in this cohort, the limited PRA information available did show higher cPRA among women than men (Supplement). Hence, the decision to bridge with tMCS may have been weighed against potential bleeding complications that may necessitate blood transfusions, as well as the risk of infectious complications if highly allosensitized individuals undergo desensitization therapies. **(14,15**) It must be noted however, that despite these potential concerns, the use of tMCS should still be considered for women meeting appropriate criteria, particularly since good outcomes can still be achieved as evidenced by a reduction in waitlist time and improved waitlist mortality in this cohort.

The IABP is the most frequently used device in both men and women listed for OHT on tMCS since it is readily accessible and has relatively fewer complications compared with ECMO or axial flow devices **(16-20)**. None-the-less, as centers gain more experience with axillary devices, such as the Impella 5.5, which provide more cardiac output and allow for more patient mobility compared to femoral IABP, practice patterns could change in the future. Trials such as DanGer Shock trial **(21)** which demonstrated a reduction in 6-month mortality with use of Impella CP among patients presenting with AMI shock may also serve as an impetus for future changes. Interestingly, despite ECMO bridge to OHT historically having poorer outcomes **(18-20)**, its use has increased significantly among both sexes post-allocation change, with a 10-fold increase among men and approximate 25-fold increase among women. Potential attributable factors may center around provider experience, improved patient selection, and higher listing status afforded to patients.

In general, the higher listing status granted by tMCS has likely contributed to reduced waitlist time for both sexes, though this is significantly more evident in women given lower BMI/BSA. Consequently, unlike other studies that have shown increased waitlist mortality in women **(5, 22)**, in our cohort, the waitlist mortality for men and women is similar and has improved post allocation presumably due to shorter wait periods and increased provider experience leading to fewer device related complications. This may have also contributed to the improvement in 1-year post-transplant survival in the post-allocation change era.

## Study Limitations

This study was a retrospective analysis and as such clinical information and outcomes were limited by the data available in the UNOS registry. Antibody sensitization data was limited with high missing rate, hence limiting analysis and conclusions regarding tMCS decision-making based on allosensitization. There may be selection bias and other confounding variables which we are unable to account for given the retrospective nature of this study.

## Conclusion

Since the UNOS allocation change, there has a been a significant increase in the use of tMCS as a bridge to OHT. Acute support devices, however, may still be underutilized in women, who account for less than a quarter of patients despite similar CI suggesting they may be just as sick as their male counterparts at listing. None-the-less, waitlist mortality has improved post allocation change for both sexes, with shorter duration on device support. One-year post transplant survival has also improved for those bridged with tMCS independent of sex.

## Data Availability

All data included in the manuscript will be made available.

## Supplement

Table showing Calculated Panel Reactive Antibody Values for Patients Listed on Temporary Mechanical Circulatory Support as Bridge to Orthotopic Heart Transplant.

|  | Pre-Allocation Change |  |  | Post Allocation Change |  |  |
| --- | --- | --- | --- | --- | --- | --- |
| Characteristic | **Women**<br>**n=203** | **Men**<br>**n=697** | **p-value** | **Women**<br>**n=1852** | **Men**<br>**n=6102** | **p-value** |
| Most recent CPRA | 18 (32) | 7 (16) | 0.002 | 20 (31) | 7 (17) | <0.001 |
| Unknown | 80 | 328 |  | 705 | 2205 |  |
| Peak CPRA | 23 (36) | 11 (21) | 0.004 | 24 (33) | 10 (20) | <0.001 |
| Unknown | 80 | 327 |  | 705 | 2209 |  |

Values are expressed as mean (SD) as appropriate. **Bold** indicates p-values that are statistically significant
CPRA = Calculated Panel Reactive Antibody

